# Using an Anomaly Detection Approach for the Segmentation of Colorectal Cancer Tumors in Whole Slide Images

**DOI:** 10.1101/2023.07.17.23292768

**Authors:** Qiangqiang Gu, Chady Meroueh, Jacob Levernier, Trynda Kroneman, Thomas Flotte, Steven Hart

**Author notes:** 200 1st St SW, Rochester, MN, 55905.

## Abstract

Colorectal cancer (CRC) is the 2^nd^ most commonly diagnosed cancer in the United States. Genetic testing is critical in assisting in the early detection of CRC and selection of individualized treatment plans, which have shown to improve the survival rate of CRC patients. The tissue slides review (TSR), a tumor tissue macro-dissection procedure, is a required pre-analytical step to perform genetic testing. Due to the subjective nature of the process, major discrepancies in CRC diagnostics by pathologists are reported, and metrics for quality are often only qualitative. Progressive context encoder anomaly detection (P-CEAD) is an anomaly detection approach to detect tumor tissue from Whole Slide Images (WSIs), since tumor tissue is by its nature, an anomaly. P-CEAD-based CRC tumor segmentation achieves a 71% ±26% sensitivity, 92% ±7% specificity, and 63% ±23% F1 score. The proposed approach provides an automated CRC tumor segmentation pipeline with a quantitatively reproducible quality compared with the conventional manual tumor segmentation procedure.

## 1. Introduction

### 1.1. Background

Colorectal cancer (CRC) is the second most frequently diagnosed cancer in the United States for both sexes and is also the second most common cause of cancer-related deaths worldwide.^1,2^ Genetic testing is the cornerstone of personalized medicine, and is rapidly becoming a necessary tool for prognostication and treatment selection, which have the potential to enhance the 5-year survival rate of CRC patients.^3^ According to the most recent NCCN guidelines,^4^ the most important factors that influence treatment selection include pathologic staging and prognostic markers, including, but not limited to, MMR status (with reflex for MLH1 promoter methylation or more expanded genomic testing), Her2 Immunostaining / Fluorescent in-situ hybridization, *KRAS, NRAS, BRAF, and NTRK* mutations. Next-generation sequencing (NGS) offers to investigate most of the above mutations / fusions.

It is important to conduct genetic testing during the diagnostic process in CRC, as 5% to 15% of cases are caused by inherited cancer susceptibility genes.^5,6^ Identifying *TP53* mutation status correlates with higher stage and influences the overall survival rate.^7^ *EGFR* inhibitor therapies are not effective for CRC patients with positive mutations in *KRAS, BRAF, PI3KCA*, and *PTEN*, highlighting the need for accurate genetic mutation status, that will ensure successful selection of individualized therapy. Different genetic mutation status also impacts CRC survival, where the CRC patients with a positive mutation of *LRP1B* have a higher recurrence rate and shorter progression-free survival (PFS) compared to those with a positive mutation of *FAT4*.^7^ Therefore, CRC genetic testing is critical in improving predictions of CRC prognostics and survival rate.

In conventional clinical CRC patient-care pathways, tumor samples are formalin-fixed and paraffin-embedded into one or more tissue blocks. A guideline by Ballester and Cruz-Correa is used to determine if individuals should undergo genetic testing based on factors such as age at diagnosis of affected family members, personal and family history of colon polyps, and extracolonic cancers.^8^ If a patient meets the guideline for genetic testing, Pathologist will confirm the best-block for testing and annotate tumor regions on H&E stained slides, to ensure selection of the largest and purest viable tumor area, followed by tumor scraping from unstained slides by cytotechnologists. This step will ensure the highest possible yield of DNA or RNA from this specimen, with least benign or inflammatory cell contaminants.

The most important factors in ensuring a successful NGS testing are preanalytical variables, including selection of invasive tumor, size of invasive tumor, viability of tumor, and purity of tumor (i.e., minimal presence of benign cells, including inflammatory cells). The current clinical workflow for NGS testing, known as tissue slide review (TSR), is completely manual and suffers from significant interindividual variation leading to discrepancies in tumor annotation.^9^ To improve on this process, we are proposing to employ an artificial intelligence (AI) tumor segmentation algorithm to automatically detect tumor regions from digitized H&E-stained whole slide images (WSIs). This would allow control of multiple pre-analytical variables through selecting the block with the largest and purest tumor surface area and segmenting that area for later tumor recovery for subsequent testing (**Figure 1**).

**Figure 1.**
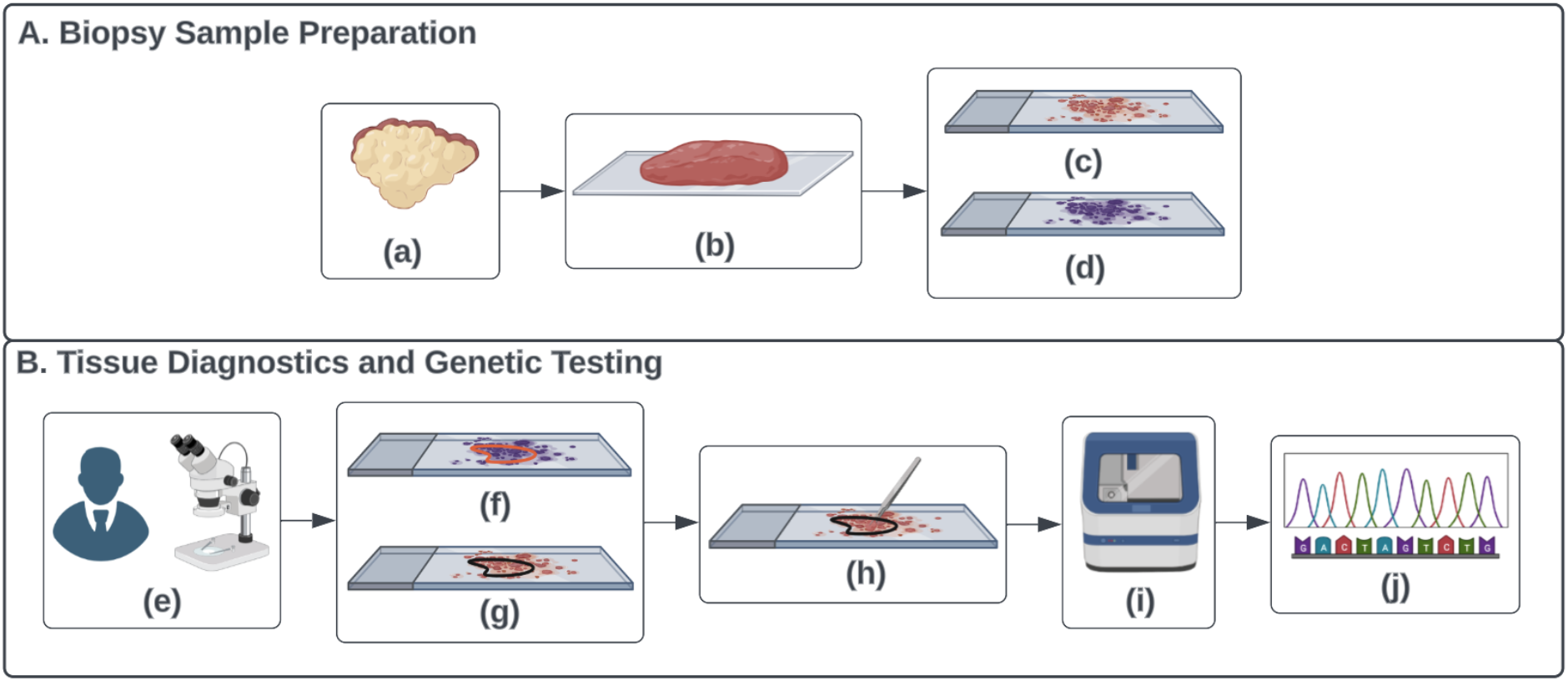
Diagram of manual workflow of tissue slide review (TSR). There are ten components included in the figure. Component (a) is a CRC tumor tissue; (b) is a cut CRC tumor biopsy sample; (c) is a glass slide with the non-stained two-dimensional CRC tumor tissue block cut from (b); (d) is a glass slide with the H&E-stained two-dimensional CRC tumor tissue block section cut from (b), which is the adjacent two-dimensional CRC tissue block section to (c); (e) illustrates the general anatomic pathology practice workflow for pathologists to make cancer diagnostics using microscope on glass slides; (f) is the pathologists diagnostics with red polygon highlighting the CRC tumor tissue regions from (d); (g) is the black CRC tumor polygon on (c) that has been aligned with the red CRC tumor polygon on (d); (h) illustrates the clinical workflow for cytotechnologists to scrape the CRC tumor tissue on (g); (i) is the NGS device used for genetic testing; (j) is the genetic testing results from the NGS technology. Two sub-figures included in this figure, A). Biopsy Sample Preparation Pipeline; B). Tissue Diagnostics and Genetic Testing Pipeline.

### 1.2. Related Work

Image classification is a widely used method for detecting tumor regions in WSIs. This approach labels a WSI as either CRC positive or negative. However, it does not provide the exact location of the tumor regions in the slide with their corresponding ***x***- and ***y***- coordinates.^10–13^ On the other hand, image segmentation provides the ***x***- and ***y***- coordinates of tumor regions in CRC WSIs - which is necessary for the TSR process (rather than a yes no answer that tumor is present).^14^ While a supervised image segmentation approach is promising, acquiring ground truth annotations from pathologists to train the supervised image segmentation model can be biased, expensive, and time consuming, making the training process impractical.

Attempting to find other approaches in order to mitigate the shortcomings of requiring pathologist-provided annotations, the use of a Generative Adversarial Network (GAN) is explored for unsupervised anomaly detection.^15^ It is used to identify patterns of pixels that deviate from the established pattern in training images, without the need for high-quality pixel-level annotations from pathologists. This approach is particularly useful in tumor segmentation, as tumor tissue is a type of anomalous colon tissue.^16,17^

The GAN-based anomaly detection algorithm, referred to as GANomaly^18^, is a commonly used unsupervised anomaly detection approach. However, the GANomaly approach is based on the deep convolutional GAN (DCGAN)^19^, which is not meant for very high-resolution images, like colon WSIs. Different from DCGAN, the progressive GAN, also known as *p*GAN, is specifically designed for high resolution image data^20^. In *p*GAN, two major components, the generator (*G*) and discriminator (*D*), are trained gradually starting from 4 × 4 resolution. Image layers of increasing resolution are incrementally added to *G* and *D*, allowing the model to be progressively trained from 4 × 4 up to 1024 × 1024, increasing by a multiple of 2, while keeping all the existing layers trainable during the entire training process. In addition, to maintain a smooth transition from lower to higher resolutions during the training of *G*, new layers are faded in smoothly while doubling the current resolution of image features using nearest neighbor filtering. A newly added *toRGB* layer with weight ***α*** increases linearly from 0 to 1, which further projects the features to the R(red)G(green)B(blue) color channels. Reversely, another newly added *fromRGB* layer with the same weight ***α*** projects the RGB color images to the feature vectors. The features are further faded into a new convolutional layer to halve their resolution using the average pooling strategy. Similarly, a smoothed training process for *D* is performed. This process could downscale the input images to match the requirements for the current image sizes of the network. This unique progressive GAN architecture is able to outperform the other conventional GAN architectures in generating photorealistic high-resolution normal colon WSIs by providing a global view focus on the normal colon histology representation from the entire slide in a relatively lower resolution level, and a local view focus on the detailed nuclei morphology patterns in a relative higher resolution level. Therefore, applying the progressive context encoder anomaly detection pipeline (P-CEAD)^21^ was proposed for CRC tumor segmentation.

## 2. Materials and Methods

The objective of this research is to automate the process of segmenting CRC tumor regions from WSI using P-CEAD. P-CEAD is a distinctive anomaly detection pipeline based on *p*GAN. Its training process consists of three phases (**Figure 2**).^**21**^ In Phase 1, a *p*GAN architecture is trained using an image inpainting technique^**22**^ on normal colon WSIs exclusively, in order to produce photorealistic normal (non-diseased) colon WSIs. This training phase enables *p*GAN to learn a reliable reference distribution of normal colon tissue representations by minimizing the error distance values between the input real normal colon WSIs and the generated photorealistic colon WSIs. Since not all pixels in a WSI are part of the tissue regions, the *Otsu*^**23**^ method was used to identify these regions and extract image patches from them. Image patches were extracted from tissue regions on WSIs in 1024 × 1024 pixels, then downsampled to 512 × 512, 256 × 256, 8 × 8, and 4 × 4 pixels. The training data is saved in TFRecord files^**24**^, with each file containing binary image patch tensors and the corresponding file name, height, width, and number of channels for each patch respective to different resolution levels. After completing phase 1 of the training, the weights of *p*GAN are frozen.

**Figure 2.**
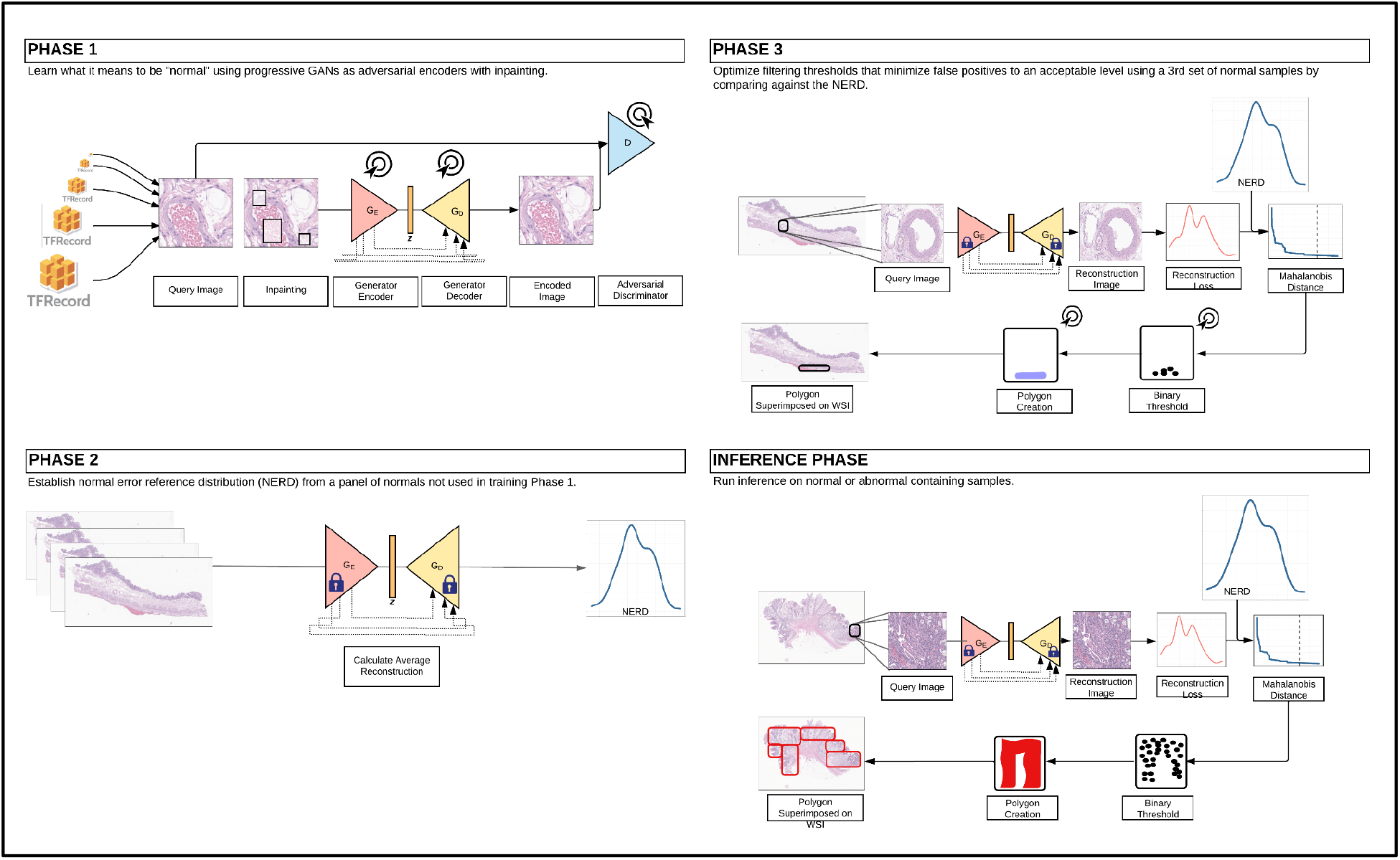
Training and Inference Pipeline Diagram of P-CEAD in CRC Tumor Segmentation. Phase 1). Phase 1 of the Training Pipeline, pGAN Training; Phase 2). Phase 2 of the Training Pipeline, Calculating NERD; Phase 3). Phase 3 of the Training Pipeline, Selecting Cut-Off Mahalanobis Distance Threshold; Inference Phase). Evaluating P-CEAD performance in CRC Tumor Segmentation.

The goal of phase 2 in the training process is to calculate the normal error reference distribution (NERD). NERD is a multivariate gaussian distribution of the absolute errors, also known as reconstruction errors, between the input real WSIs and the generated photorealistic WSIs. Because, during phase 1, *p*GAN is only trained on normal colon WSIs, the absolute errors between the input real normal colon WSIs and the generated photorealistic normal colon WSIs should be small. The reconstruction errors between the input real CRC and the generated photorealistic CRC WSIs are expected to be relatively large because the GAN never learned how to encode features present in anomalous tissues and is therefore more prone to create higher reconstruction errors.

During phase 3 of the training, the NERD and reconstruction errors are used to calculate pixel-level Mahalanobis distances. The goal of this phase is to identify a cut-off threshold to distinguish between normal and CRC tumor pixels in a WSI. If the Mahalanobis distance for a given pixel is higher than the threshold, it is considered an abnormal colon pixel; otherwise, it is considered a normal colon pixel.

After completing all three phases of training, the *p*GAN model was fed 1024 × 1024 resolution image patches extracted from tissue regions on a test set of WSI containing CRC. From this, the reconstruction errors between the input and generated images from the trained *p*GAN were calculated and binarized based on the Mahalanobis distance threshold. Using the *shapely* package for Python^25^, polygon objects were created around the identified CRC tumor pixels and saved into a GeoPandas dataframe^25^. The comparison between predicted and pathologist-annotated CRC tumor polygon objects were used to calculate a confusion matrix, including pixel-level counts of true positive (TP), false positive (FP), true negative (TN), and false negative (FN) areas of the WSI. TP was defined as the number of pixels of the areas that are within both the predicted and annotated CRC tumor polygons. FP was defined as the number of pixels of the areas that are within the predicted CRC tumor polygons but are not within the annotated CRC tumor polygons. TN was defined as the number of pixels of the areas that are outside of both the predicted and annotated CRC tumor polygons. FN was defined as the number of pixels of the areas that are outside of the predicted CRC tumor polygons but are within the annotated CRC tumor polygons. Sensitivity, specificity, and accuracy are derived from these values to provide a quantitative measurement of the model performance. The codebase, including the training and inference pipeline, is publicly available via https://github.com/quincy-125/tsr_crc_tumor_seg.

A total of 277 WSIs scanned by the Aperio GT450 scanner^26^ at the Mayo Clinic were used for training and inference (Table 1). Out of these, 140 were normal colon WSIs and 137 were CRC WSIs. All WSIs underwent quality control examination by a senior cytotechnologist and a senior anatomic pathologist. During the training process, 140 normal colon WSIs were used. Out of these, 100 were used for Phase 1, 20 were used for Phase 2, and the remaining 20 were used for Phase 3. Model inference was performed using all 137 CRC WSIs. The manual annotations of CRC tumors were required to compute the statistical metrics (i..e, confusion matrix, sensitivity, specificity, and accuracy). Tumor annotations from all 137 CRC WSIs were drawn by pathologists using QuPath.^27^

**Table 1.** Data Information Summary Table with WSI Type and Number of WSIs Information Regarding Each of the Three Training Phases and One Inference Phase.

| Phase Name | Slide Type | Number of Slides |
| --- | --- | --- |
| Training Phase 1 | Normal Colon | 100 |
| Training Phase 2 | Normal Colon | 20 |
| Training Phase 3 | Normal Colon | 20 |
| Inference Phase | Colorectal Cancer | 137 |

## 3. Results & Discussion

The P-CEAD model demonstrated a sensitivity, specificity, and accuracy of 71% ± 26%, 92% ± 7%, and 63% ± 23%, respectively, as determined by the confusion matrix values derived from the inference results of 137 CRC tumor WSI (**Figure 3**).

**Figure 3.**
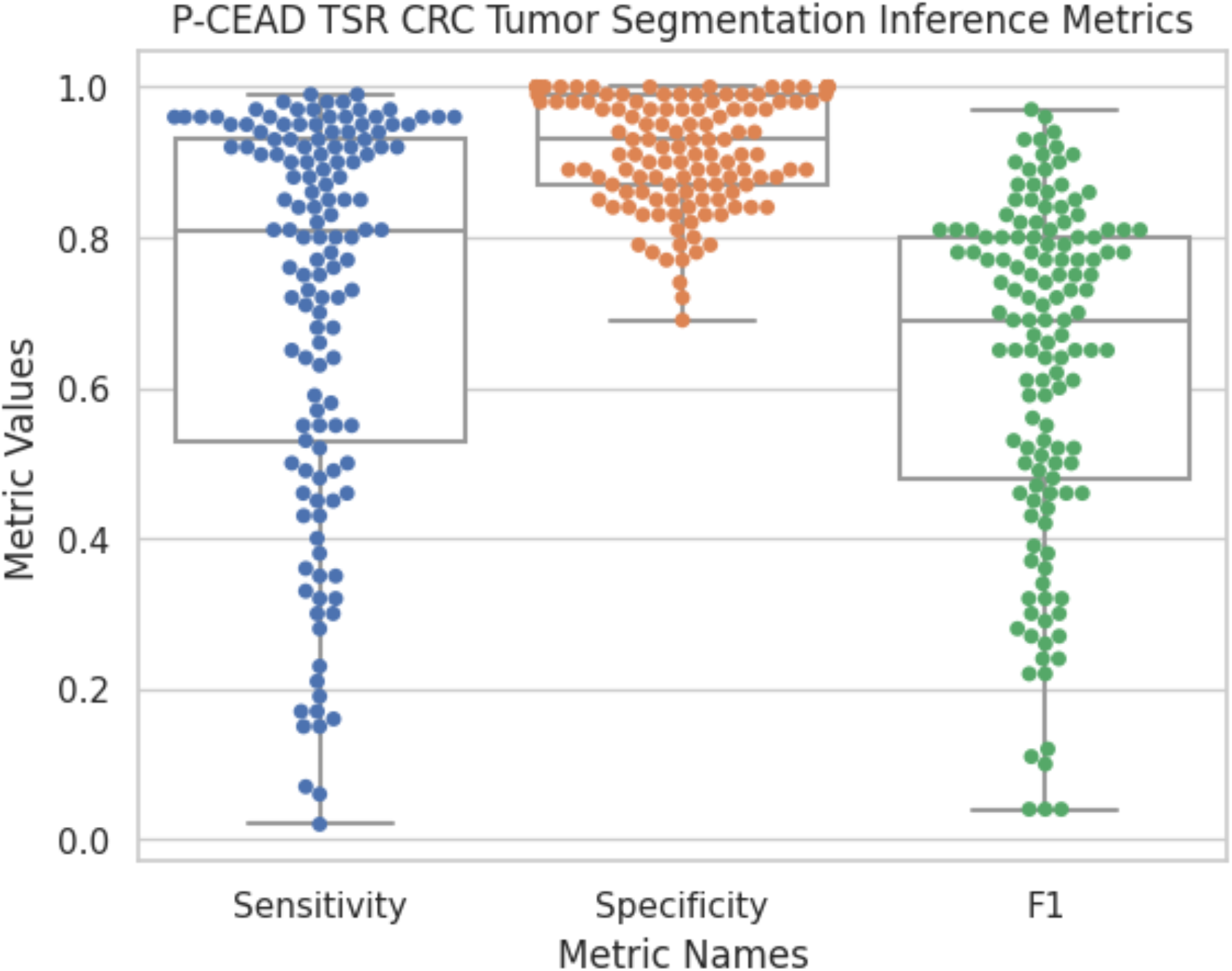
Quantitative Measurement Results of P-CEAD Inference Performance in CRC Tumor Segmentation on 137 CRC Tumor WSIs. The Statistical Metrics Including the Sensitivity, Specificity, and F1 Score. Each CRC WSI is a blue dot.

A notable advantage of the P-CEAD-based CRC tumor segmentation pipeline is its fully unsupervised nature. This eliminates the need for time-consuming and costly pathologist annotations during the training process, underlining one of the benefits of implementing the unsupervised P-CEAD approach for CRC tumor segmentation in WSI.

However, our ground truth for CRC tumor annotation, meticulously done by a pathologist, is microscopic and encapsulates large non-tumor areas surrounding the main lesion, inclusive of whitespace regions. These anomalies are a source of model error since our model focuses only on tissue-containing patches, excluding the whitespace regions. Consequently, the manually annotated CRC tumor areas (TP) tend to be larger than the predicted tumor areas (TP+FP), leading to an increase in false negative predictions **(Figure 4 A**). One potential solution could be to remove whitespace regions from the manually annotated areas to reduce the false negatives in future iterations.

**Figure 4.**
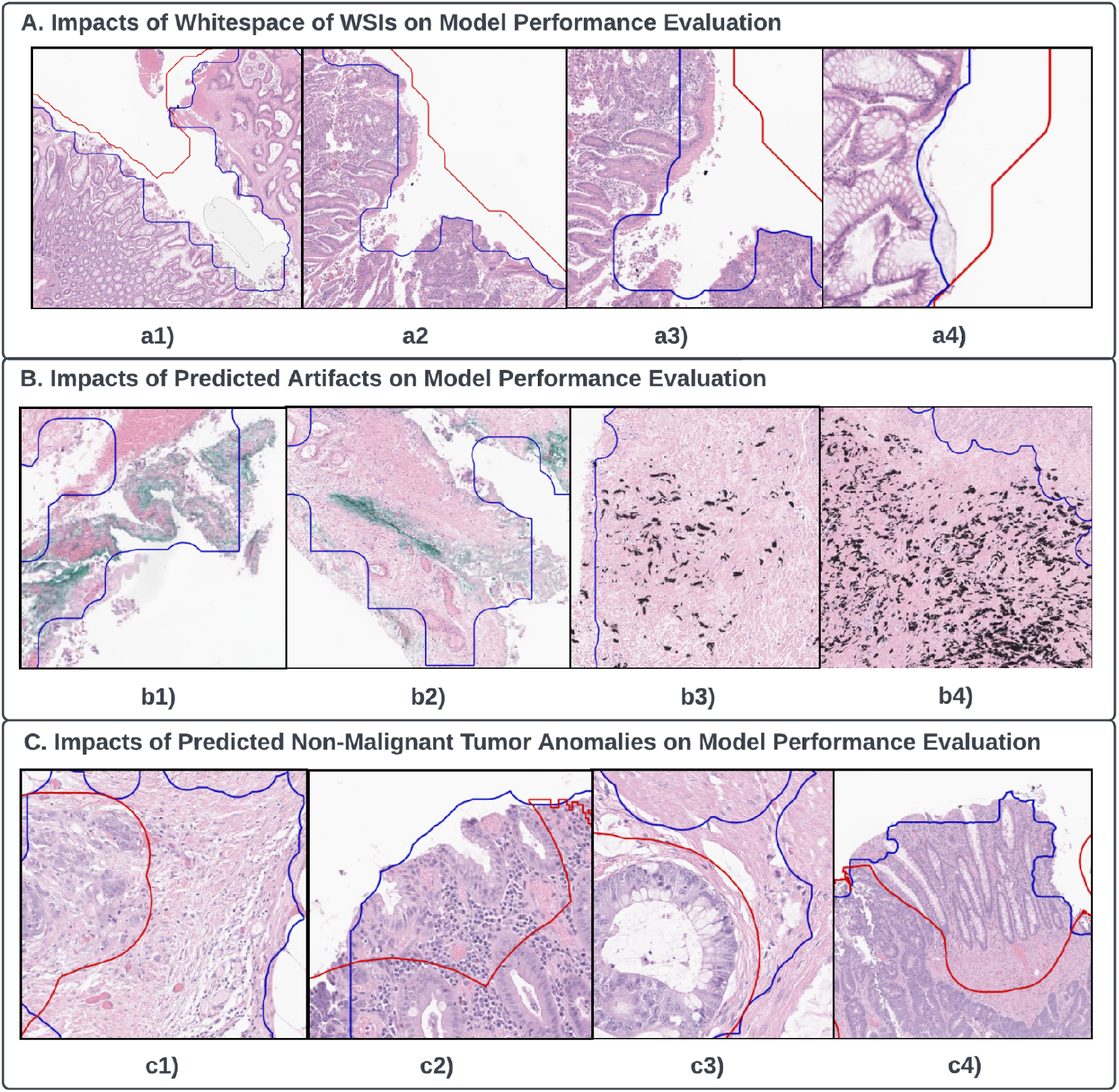
Qualitative Model Performance Evaluations. Note that the red lines are the boundaries of the ground truth CRC tumor annotations; the blue lines are the boundaries of the predicted CRC tumor areas. A). Impacts of whitespace of WSIs on model performance evaluation with a1) - a4) four example patches. All whitespace areas presented on a1) - a4) are all included in manual CRC tumor annotation regions, but not included in the model prediction regions. B). Impacts of predicted artifacts on model performance evaluation with b1) - b4) four example patches. b1) and b2) are example patches with green on-slide annotation inks that are within the model prediction regions, but outside the manual CRC tumor annotation regions. b3) and b4) are example patches with black on-slide annotation inks that are within the model prediction regions, but outside the manual CRC tumor annotation regions. C). Impacts of predicted non-malignant CRC tumor anomalous tissue on model performance evaluation with c1) - c4) four example patches. On each of the two example patches, c1) and c3), tissues on the left to the red polygon boundary line are included in the manual CRC tumor annotations; tissues on the right to the red polygon boundary line are not included in the manual CRC tumor annotations but included in the model prediction regions. On each of the rest two example patches, c2) and c4), tissues on the lower regions toward the red boundary line are included in both the annotated and predicted CRC tumor areas; tissues on the upper regions toward the red boundary line are only included in the predicted- but are not included in the annotated CRC tumor areas.

Originally designed as an anomaly detection model, P-CEAD identifies all regions diverging from the norm, which includes inked tissue, inflamed tissue, and malignant areas from WSI. This could lead the model to classify artifacts such as on-slide annotations as anomalies, thereby increasing the false positive predictions (Figure 4 B). To mitigate this, we propose adopting Jiang et al.’s^28^ ink-removal technique as part of the data preprocessing procedure before model inference in future experiments.

In our P-CEAD-based model, peritumoral changes were included in the predicted CRC tumor areas. As discussed earlier, P-CEAD aims to detect all anomalous tissues, not solely malignant CRC tumors. Hence, the model included benign stromal tissue connected to malignant CRC tumors within the predicted areas, a factor contributing to false positives. For model training, we relied on normal colon WSIs (Section 2). A potential amendment could be to introduce benign tissues into the training set to adjust the Normalized Error Rate Difference (NERD), thereby reducing false positive predictions from non-malignant tissues (**Figure 4 C**)

In summary, our P-CEAD model, an unsupervised anomaly detection-based tumor segmentation approach, yielded 71% ± 26% sensitivity, 92% ± 7% specificity, and 90% ± 7% accuracy in segmenting CRC tumors from WSI. This underscores the value in further exploration of the P-CEAD-based tumor segmentation algorithm in other cancer types. To optimize model performance, we recommend adding WSIs with artifacts or non-malignant CRC tumor anomalous tissue to the training data set. This could reduce the misclassification of such tissues as malignant CRC tumors when utilizing the anomaly detection approach of P-CEAD. Further, image preprocessing approaches such as ink-removal and whitespace removal using the Otsu method could enhance both quantitative (i.e., reducing false positives and negatives) and qualitative model performance.

## Data Availability

All data produced in the present study are available upon reasonable request to the authors

## Funding

This work was supported by the Mayo Clinic Digital Pathology Program, and the University of Minnesota Graduate School Doctoral Dissertation Fellowship for the year of 2022-2023.

## Acknowledgements

QG, JL implemented the training and inference pipeline of P-CEAD and did all the experiments. CM, TK, and TF provided the data, and annotations for tumor ground-truth. TF and SH oversaw the project.

## Declaration of Interests

The authors declare that they have no known competing financial interests or personal relationships that could have appeared to influence the work reported in this paper.

## Declaration of Generative AI in Scientific Writing

During the preparation of this work the author(s) used ChatGPT in order to assist language editing in the writing process. After using this tool/service, the author(s) reviewed and edited the content as needed and take(s) full responsibility for the content of the publication.

## Notes

### Competing Interest Statement

The authors have declared no competing interest.

### Funding Statement

This work was funded by the Mayo Clinic Digital Pathology Program, and the University of Minnesota Graduate School Doctoral Dissertation Fellowship for the year of 2022-2023.

### Author Declarations

The Institutional Review Board of the Mayo Clinic gave approval of this study.

## Reference

1. Colorectal cancer: Epidemiology, risk factors, and protective factors. Accessed May 30, 2023. https://www.medilib.ir/uptodate/show/2606

2. Bray F, Ferlay J, Soerjomataram I, Siegel RL, Torre LA, Jemal A. Global cancer statistics 2018: GLOBOCAN estimates of incidence and mortality worldwide for 36 cancers in 185 countries. CA Cancer J Clin. 2018;2018(6):394–424. doi:10.3322/caac.21492

3. Dulskas A, Gaizauskas V, Kildusiene I, Samalavicius NE, Smailyte G. Improvement of Survival over Time for Colorectal Cancer Patients: A Population-Based Study. J Clin Med. 2020;2020(12):4038. doi:10.3390/jcm9124038

4. Permission to Cite or Use NCCN Content. NCCN. Accessed May 31, 2023. https://www.nccn.org/guidelines/permission-to-cite-or-use-nccn-content

5. Uson PLS, Riegert-Johnson D, Boardman L, et al. Germline Cancer Susceptibility Gene Testing in Unselected Patients With Colorectal Adenocarcinoma: A Multicenter Prospective Study. Clin Gastroenterol Hepatol Off Clin Pract J Am Gastroenterol Assoc. 2022;2022(3):e508–e528. doi:10.1016/j.cgh.2021.04.013

6. Moretz C, Byfield SD, Hatchell KE, et al. Comparison of Germline Genetic Testing Before and After a Medical Policy Covering Universal Testing Among Patients With Colorectal Cancer. JAMA Netw Open. 2022;2022(10):e2238167. doi:10.1001/jamanetworkopen.2022.38167

7. Zhuang Y, Wang H, Jiang D, et al. Multi gene mutation signatures in colorectal cancer patients: predict for the diagnosis, pathological classification, staging and prognosis. BMC Cancer. 2021;2021(1):380. doi:10.1186/s12885-021-08108-9

8. Ballester V, Cruz-Correa M. How and When to Consider Genetic Testing for Colon Cancer? Gastroenterology. 2018;2018(4):955–959. doi:10.1053/j.gastro.2018.08.031

9. Smits LJH, Vink-Börger E, van Lijnschoten G, et al. Diagnostic variability in the histopathological assessment of advanced colorectal adenomas and early colorectal cancer in a screening population. Histopathology. 2022;2022(5):790–798. doi:10.1111/his.14601

10. Sari CT, Gunduz-Demir C. Unsupervised Feature Extraction via Deep Learning for Histopathological Classification of Colon Tissue Images. IEEE Trans Med Imaging. 2019;2019(5):1139–1149. doi:10.1109/TMI.2018.2879369

11. Boyd J, Liashuha M, Deutsch E, Paragios N, Christodoulidis S, Vakalopoulou M. Self-Supervised Representation Learning using Visual Field Expansion on Digital Pathology. Published online September 7, 2021. doi:10.48550/arXiv.2109.03299

12. Neto PC, Oliveira SP, Montezuma D, et al. iMIL4PATH: A Semi-Supervised Interpretable Approach for Colorectal Whole-Slide Images. Cancers. 2022;2022(10):2489. doi:10.3390/cancers14102489

13. Wang KS, Yu G, Xu C, et al. Accurate diagnosis of colorectal cancer based on histopathology images using artificial intelligence. BMC Med. 2021;19:76. doi:10.1186/s12916-021-01942-5

14. Deep learning-based histopathological segmentation for whole slide images of colorectal cancer in a compressed domain | Scientific Reports. Accessed May 30, 2023. https://www.nature.com/articles/s41598-021-01905-z

15. Goodfellow IJ, Pouget-Abadie J, Mirza M, et al. Generative Adversarial Networks. Published online June 10, 2014. doi:10.48550/arXiv.1406.2661

16. Goldstein M, Uchida S. A Comparative Evaluation of Unsupervised Anomaly Detection Algorithms for Multivariate Data. PLOS ONE. 2016;2016(4):e0152173. doi:10.1371/journal.pone.0152173

17. Lee Y, Kang P. AnoViT: Unsupervised Anomaly Detection and Localization with Vision Transformer-based Encoder-Decoder. Published online March 21, 2022. doi:10.48550/arXiv.2203.10808

18. Akcay S, Atapour-Abarghouei A, Breckon TP. GANomaly: Semi-Supervised Anomaly Detection via Adversarial Training. Published online November 13, 2018. doi:10.48550/arXiv.1805.06725

19. Radford A, Metz L, Chintala S. Unsupervised Representation Learning with Deep Convolutional Generative Adversarial Networks. Published online January 7, 2016. doi:10.48550/arXiv.1511.06434

20. Karras T, Aila T, Laine S, Lehtinen J. Progressive Growing of GANs for Improved Quality, Stability, and Variation. Published online February 26, 2018. doi:10.48550/arXiv.1710.10196

21. Using Progressive Context Encoders for Anomaly Detection in Digital Pathology Images | bioRxiv. Accessed May 30, 2023. https://www.biorxiv.org/content/10.1101/2021.07.02.450957v1.full

22. Jam J, Kendrick C, Drouard V, Walker K, Hsu GS, Yap MH. Symmetric Skip Connection Wasserstein GAN for High-Resolution Facial Image Inpainting. Published online September 12, 2020. doi:10.48550/arXiv.2001.03725

23. Otsu N. A Threshold Selection Method from Gray-Level Histograms. IEEE Trans Syst Man Cybern. 1979;1979(1):62–66. doi:10.1109/TSMC.1979.4310076

24. Abadi M, Barham P, Chen J, et al. TensorFlow: A system for large-scale machine learning. Published online May 31, 2016. doi:10.48550/arXiv.1605.08695

25. GeoPandas 0.13.0 — GeoPandas 0.13.0+0.gaa5abc3.dirty documentation. Accessed May 30, 2023. https://geopandas.org/en/stable/

26. Aperio GT 450 - Automated, High Capacity Digital Pathology Scanner. Accessed May 30, 2023. https://www.leicabiosystems.com/us/digital-pathology/scan/aperio-gt-450/

27. Bankhead P, Loughrey MB, Fernández JA, et al. QuPath: Open source software for digital pathology image analysis. Sci Rep. 2017;7:16878. doi:10.1038/s41598-017-17204-5

28. Jiang J, Prodduturi N, Chen D, et al. Image-to-image translation for automatic ink removal in whole slide images. J Med Imaging Bellingham Wash. 2020;2020(5):057502. doi:10.1117/1.JMI.7.5.057502

